# Environmental reservoirs of high-risk ESBL- and carbapenemase-producing *E. coli* and *Klebsiella* in maternity wards in Yaounde (Cameroon): Whole-genome sequencing and antimicrobial susceptibility studies

**DOI:** 10.64898/2026.03.16.26348525

**Authors:** Gabriel Cedric Bessala, Germanie Delaisie Abomo, Rodrigue Ngamaleu, Felix Essiben, Nicole Wheeler, Michelle M. C. Buckner, Jan-Ulrich Kreft, Blaise Pascal Bougnom

**Author notes:** **Correspondence: Blaise Pascal Bougnom**.

## Abstract

**Background:** The hospital environment is increasingly recognized as a critical reservoir for antimicrobial-resistant (AMR) bacteria. In sub-Saharan Africa, maternity wards represent high-risk settings where environmental contamination poses a direct threat to vulnerable mothers and neonates. Despite this, there is a significant lack of data integrating phenotypic resistance with whole-genome sequencing (WGS) to understand antimicrobial resistance (AMR) in these settings. This study characterized the AMR patterns and genomic features of ESBL-producing *Escherichia coli* and *Klebsiella* spp. isolated from maternity ward surfaces in Yaounde, Cameroon.

**Methods:** A cross-sectional environmental study was conducted across four maternity wards. Isolates were identified via standard microbiological methods, and antimicrobial susceptibility testing against 13 antibiotics was performed following EUCAST 2024 guidelines. Short-read WGS was utilized to identify sequence types (STs), plasmid incompatibility groups, antibiotic resistance genes (ARGs), and virulence factors. Plasmid‒ARG association networks were constructed to visualize resistance dynamics.

**Results:** Nineteen ESBL-producing *Enterobacterales* were identified, comprising 15 *E. coli* and four *Klebsiella* isolates. High levels of multidrug resistance were observed against ciprofloxacin, penicillins, and third-generation cephalosporins. While the isolates remained sensitive to colistin and imipenem, alarming resistance to meropenem was detected. Genomic analysis revealed the presence of globally disseminated high-risk lineages, including *E. coli* ST131, ST1193, and ST410, alongside *Klebsiella* ST1324 and ST489. Critical resistance determinants, including ESBLs, AmpC enzymes, and carbapenemases (NDM and OXA-48-like), are frequently associated with epidemic plasmids such as IncF, IncA/C2, and IncL/M. Additionally, the isolates harboured virulence factors characteristic of extraintestinal pathogenic *Enterobacterales*.

**Conclusions:** The widespread presence of high-risk carbapenemase-producing clones on maternity ward surfaces identifies the hospital environment as a significant AMR reservoir in Yaounde. These findings highlight the urgent need for reinforced infection prevention and control (IPC) measures, robust antimicrobial stewardship, and the integration of genomic surveillance to safeguard highly susceptible maternal and neonatal populations from life-threatening infections.

## Introduction

Neonatal and maternal mortality are major health challenges facing Cameroon and other sub-Saharan African countries. Recent data indicate that the neonatal mortality rate in Cameroon is approximately 28 deaths per 1,000 live births [1]. While global maternal mortality has declined, the ratio remains high in Cameroon, with approximately 529 deaths per 100,000 live births [2].

Neonatal infections such as sepsis, pneumonia, and healthcare-associated infections (HAIs) have been identified as the leading causes of neonatal death in low- and middle-income countries (LMICs) [3]. Consistent reports from sub-Saharan Africa show that maternal and neonatal sepsis are among the top causes of death, emphasizing the need to strengthen infection prevention in maternity and neonatal units [3,4]. Such epidemiological data highlight the importance of examining infection risks in maternity wards, where highly susceptible mothers and newborns are exposed to hospital environments during their most delicate times of care.

HAIs significantly cause sickness and death and increase healthcare costs worldwide, but the burden is disproportionately greater in LMICs where there is limited infection prevention and control (IPC) infrastructure, overcrowding, and insufficient surveillance [5,6]. Pathogens of concern include extended-spectrum beta-lactamase (ESBL)-producing *Escherichia coli* and *Klebsiella pneumoniae*, which are considered critical priority pathogens by the World Health Organization because of their multidrug resistance and role in severe invasive infections [7]. In Africa, these pathogens are frequently the cause of maternal and neonatal sepsis, as antimicrobial resistance (AMR) levels rise rapidly [8].

The hospital setting has been increasingly studied as a major reservoir and transmission route for AMR. Contaminated surfaces, medical devices, and healthcare workers’ hands play a role in the survival and indirect spread of multidrug-resistant bacteria [9]. Environmental contamination has been linked to the spread of ESBL-producing *Enterobacterales* in high-risk units such as intensive care and neonatal wards [10]. However, evidence from African hospitals remains limited and disjointed, especially in maternity settings.

Maternity wards represent some of the most underresearched areas in LMICs despite their susceptibility to HAIs due to high patient turnover and numerous invasive procedures [11]. Studies in sub-Saharan Africa have revealed ESBL-producing *E. coli* and *Klebsiella* spp. in hospital clinical isolates, including those from neonatal and obstetric units [12,13]. Despite this, very few comprehensive studies have integrated environmental sampling, phenotypic resistance profiling, and genomic characterization in these settings. Whole-genome sequencing (WGS) allows for the mapping of resistance genes and transmission mechanisms at a fine scale, but its usage remains largely unavailable in LMIC maternity wards [14].

Thus, this investigation determined the extent of environmental contamination in maternity wards in Yaounde, Cameroon, by ESBL-producing *E. coli* and *Klebsiella* spp. By utilizing extensive environmental sampling and WGS, this work helps narrow the evidence gap concerning the environmental sources of AMR to reinforce data-driven IPC and antimicrobial stewardship (AMS) interventions.

## Materials and methods

### Study design and setting

The study was carried out in Yaoundé, the political capital of Cameroon. The city has more than four million inhabitants. Maternity wards in the following hospitals were investigated: Yaounde Central Hospital (HCY) in the Yaounde I district, Cite Verte district hospital (HDC) in the Yaounde V district, Nkolndongo district hospital (HDN) in the Yaounde III district, and Odza district hospital (HDO) in the Yaounde VI district. These healthcare facilities were chosen because they cover different types of wards found across diverse administrative districts of the city.

Sample collection

Environmental samples from the surfaces and air of the four maternity wards were collected monthly from January to November 2024. Sampling targeted nine functional areas that are commonly involved in patient care and staff activities, including (1) delivery rooms, (2) postpartum wards, (3) neonatal care units, (4) consultation rooms, (5) operating theaters, (6) corridors, (7) waiting areas, (8) sanitary facilities, and (9) waste zones. These areas were selected on the basis of their high frequency of human contact, potential microbial contamination, and association with IPC practices. To ensure the representativeness of the results, sampling was executed during routine clinical activity. All sampling material-handling techniques were aseptic, and in each session, field controls were taken to keep track of possible external contamination related to handling, transport, or background environmental sources.

Surface sampling was performed via sterile swabs premoistened with buffered peptone water. Swabbing was carried out on high-touch and high-risk surfaces such as bed rails, delivery tables, incubators, medical equipment, door handles, sinks, worktops, and waste container lids. Each surface was swabbed over an approximate area of 10 cm × 10 cm via a standardized multidirectional technique to maximize the recovery of microorganisms. After surface sampling, the swabs were immediately placed in sterile transport tubes.

Air sampling was conducted in delivery rooms, postpartum wards, and neonatal care units to assess airborne microbial contamination. Passive air sampling followed standard settling-plate procedures in which MacConkey agar plates supplemented with cefotaxime (2 mg/L) were exposed for 1 h at a height of 1 meter above the floor and at least 1 meter away from walls or obstacles [15]. Following exposure, the plates were sealed and labelled. Both swabs and plates were transported within four hours of collection in cool boxes (4–8°C) to the laboratory for processing.

Isolation and identification of ESBL-producing *E. coli* and *Klebsiella* spp.

All the environmental surface and air samples were immediately processed upon arrival at the laboratory. The surface swabs were streaked onto MacConkey agar supplemented with 2 mg/L cefotaxime as a selective medium for the isolation of cefotaxime-resistant gram-negative enteric bacteria. The plates harboring the air samples and streaked swabs were incubated at 37°C for 24 hours, after which colonies showing the typical morphology of *E. coli* or *Klebsiella* spp. and fermented lactose were selected for further identification. When more than one colony morphotype was present on a plate, representative colonies were purified to recover different resistant strains. Presumptive isolates were streaked onto CHROMagar™ ESBL medium (CHROMagar, Paris, France) to confirm resistance and provide preliminary species identification on the basis of colony color from hydrolysis of chromogenic substrates. All colonies with a colouration consistent with those specified in the manufacturer’s instruction leaflet to be indicative of ESBL-producing *E. coli* or *Klebsiella* spp. were subcultured until purity and identified via biochemical methods via the API 20E system (bioMérieux, Marcy-l’Etoile, France). The identification results were interpreted via the API database. All the isolates confirmed as ESBL-producing *E. coli* or *Klebsiella* spp. were stored in 20% glycerol stocks at - 80°C for future antimicrobial susceptibility testing and WGS.

### Antimicrobial susceptibility testing (AST)

The antimicrobial susceptibility of all ESBL-producing *E. coli* and *Klebsiella* was determined via the Kirby–Bauer disk diffusion method on Mueller‒Hinton agar (MHA) in accordance with EUCAST 2024 recommendations [16]. *Escherichia coli* ATCC 25922 and *Klebsiella pneumoniae* ATCC 700603 served as quality control microorganisms. The turbidity of the bacterial suspensions was adjusted to the turbidity of a 0.5 McFarland standard; sterile swabs were used to inoculate MHA plates uniformly. The following antimicrobials were evaluated (per mL): ampicillin 10 µg, amoxicillin-clavulanic acid 20/10 µg, cefotaxime 30 µg, ceftazidime 30 µg, cefepime 30 µg, cefoxitin 30 µg, aztreonam 30 µg, ciprofloxacin 5 µg, levofloxacin 5 µg, gentamicin 10 µg, amikacin 30 µg, tetracycline 30 µg, trimethoprim-sulfamethoxazole 1.25/23.75 µg, chloramphenicol 30 µg, imipenem 10 µg, meropenem 10 µg, and ertapenem 10 µg. The antibiogram results were interpreted as resistant (R), susceptible (S), or “susceptible at increased exposure (I) according to the EUCAST 2024 breakpoints.

### DNA extraction, library preparation, and whole-genome sequencing

DNA extraction, library preparation and WGS of all the ESBL-producing *Escherichia coli* and *Klebsiella* isolates were conducted at MicrobesNG, Birmingham, UK, on an Illumina HiSeq 2500 platform to generate 250 bp paired-end reads (min. 30x coverage per genome).

### Bioinformatics and genomic analysis

Raw sequence reads were processed through the standardized bioinformatics pipeline of MicrobesNG, comprising an initial quality check of FASTQ files via FastQC v0.11.9 and removing low-quality bases together with adapter sequences via Trimmomatic v0.39 [17]. The cleaned reads were de novo assembled via the SPAdes v3.15.5 assembler with default parameters [18]. The assembly quality (N50, total length, and number of contigs) was checked via QUAST v5.2.0 [19]. Contigs shorter than 500 bp or with coverage less than 10× were removed from further analysis. Species confirmation was achieved via Mash and Kraken2, whereas MLST was performed via the mlst tool [20] with the PubMLST database (https://pubmlst.org/). Plasmid replicons were detected in silico via PlasmidFinder, with thresholds set to ≥95% identity and ≥60% coverage [21]. Antibiotic resistance- and virulence-associated genes were detected via ABRicate against the ResFinder v4.7.2, CARD v3.3.0, ARG-ANNOT v6, and VFDB v6.0 databases with settings of ≥90% identity and ≥90% coverage [22]. For phylogenomic analysis, the genomes were initially annotated with Prokka v1.14.6 [23]. Then, the pangenome analysis software Roary v3.13.0 [24] was used, with a ≥95% identity threshold for BLASTp, to generate the core-genome alignment for phylogeny. Maximum likelihood trees were generated with IQ-TREE v2.1.4 [25], with the best-suited nucleotide substitution model identified with ModelFinder. Support values were obtained with ultrafast bootstrapping with 1,000 replicates. The trees were visualized and annotated via iTOL v6 [26].

## Results

### Environmental isolation and species distribution

From the 1,519 environmental samples taken, including 231 air samples and surface swabs, nineteen ESBL-producing *E. coli* and *Klebsiella* isolates were obtained from surfaces in the four investigated maternity wards in Yaounde, Cameroon. The air-exposed plates did not yield any ESBL-producing *E. coli* or *Klebsiella* spp. These strains consisted of fifteen isolates of *E. coli* and four *Klebsiella* spp., including two *Klebsiella pneumoniae* and two *Klebsiella quasipneumoniae* isolates. The highest number of isolates (n=12; ten *E. coli* and two *Klebsiella pneumoniae*) were found at HCY, followed by HDO (n = 4; three *E. coli* and one *Klebsiella pneumoniae*), HDCV (n = 3; three *E. coli*), and HDN (n = 2; one *E. coli* and one *Klebsiella pneumoniae*) (Figure 1).

**Figure 1.**
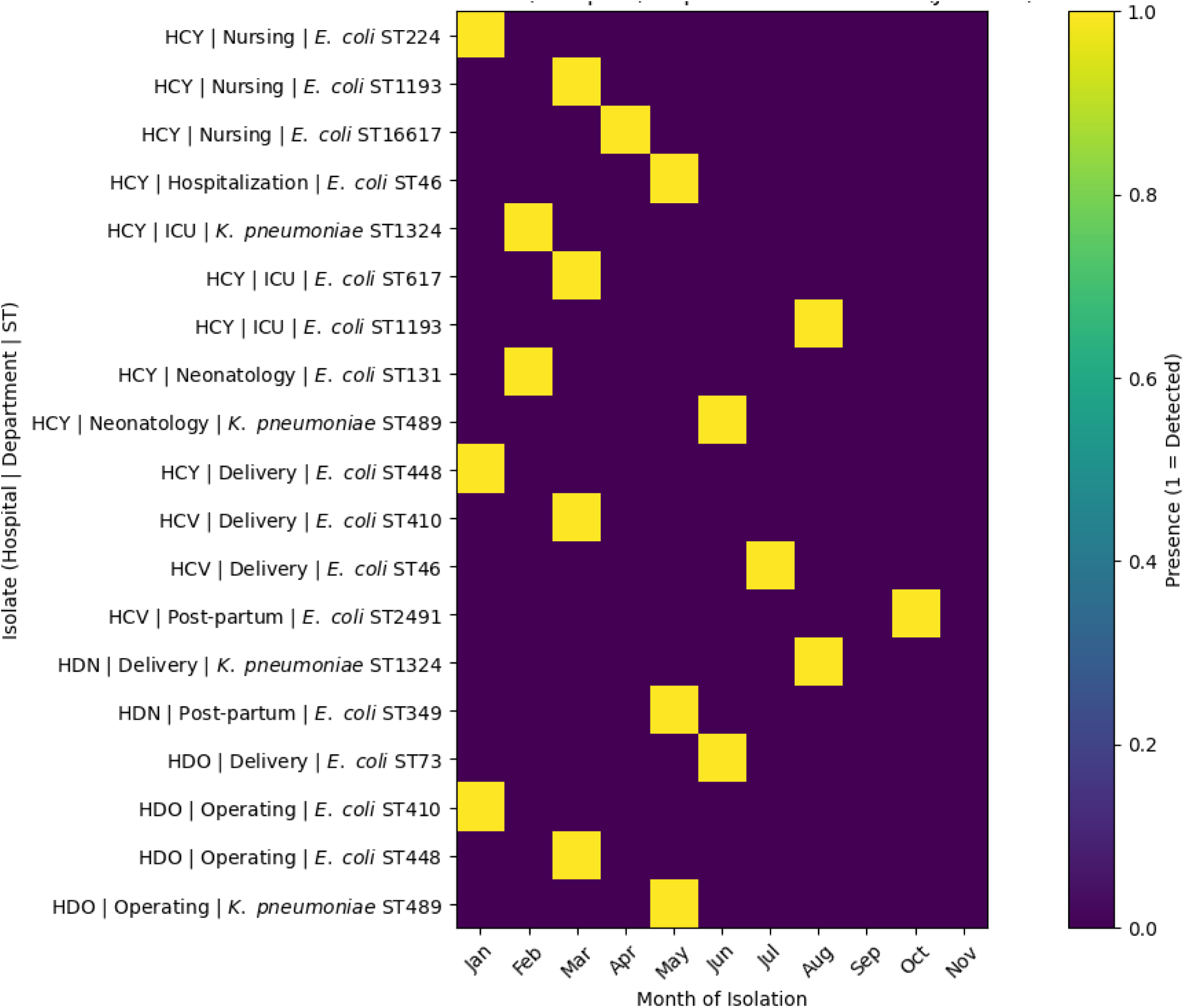
Heatmap showing monthly distribution of ESBL-producing *E. coli* and *K. pneumoniae* across hospitals and maternity departments.

From a spatial perspective, isolates were predominantly isolated from high-contact maternity environments such as intensive care, neonatology, delivery wards, hospitalization units, and operating theatres, whereas observation areas, offices, and several postpartum and newborn care units did not yield any isolates. From a temporal standpoint, ESBL-producing isolates were detected sporadically during the study period, with the continuous presence of clinically significant *E. coli* STs (ST131, ST410, ST617, ST1193, ST73, and ST448) and *K. pneumoniae* strains ST1324 and ST489 in several departments. In summary, ESBL-producing *E. coli* and *Klebsiella* spp. were mainly found on surfaces and were more commonly detected at tertiary facilities (HCYs) than at district hospitals.

### Antimicrobial susceptibility profiles

All nineteen isolates had an MDR phenotype (Table S1) and were resistant to at least eight of the thirteen antibiotics used, which covered four drug classes. Overall, 18 out of 19 strains were resistant to ten or more antibiotics. All the strains were resistant to amoxicillin, amoxicillin-clavulanic acid and ciprofloxacin. There is universal resistance to third-generation cephalosporins, including ceftriaxone, ceftazidime, and cefotaxime, as well as cefoxitin. Resistance to aztreonam occurred in sixteen isolates, while three isolates were susceptible. Testing for carbapenem susceptibility revealed a disparity between the two antibiotics evaluated. All the isolates were resistant to meropenem but susceptible to imipenem. Many isolates were resistant to aminoglycosides: tobramycin resistance was observed in fifteen isolates, whereas gentamicin resistance was found in eleven isolates. Six isolates were sensitive to gentamicin, whereas two isolates were susceptible to increased exposure. Sensitivity to colistin was maintained in eighteen isolates, whereas resistance was observed in one isolate. The susceptibility profiles of all the isolates are summarized in a heatmap (Figure 2).

**Figure 2.**
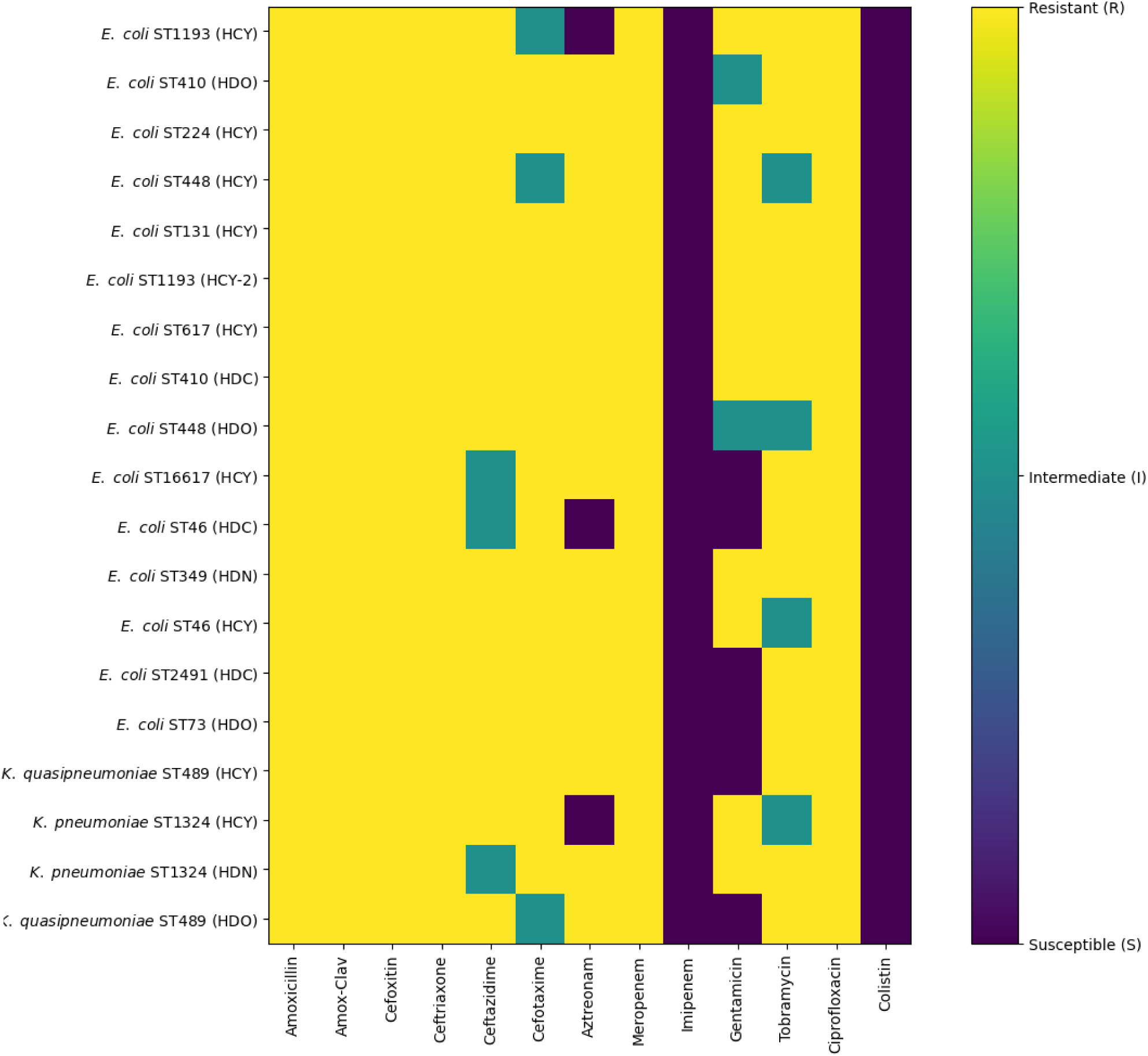
Heatmap of antimicrobial susceptibility profiles of ESBL-producing *Escherichia coli* and *Klebsiella* isolates recovered from environmental surfaces of maternity wards in Yaoundé, Cameroon. Susceptibility results are shown for β-lactams, aminoglycosides, fluoroquinolones, and polymyxins. Colors represent resistant (R), intermediate (I), and susceptible (S) phenotypes.

### Sequence types of *Escherichia coli* and *Klebsiella* species

MLST analysis revealed that *E. coli* and *Klebsiella* isolates from maternity ward surfaces presented high genetic diversity, and more than one *E. coli* ST, as well as *Klebsiella* ST, coexisted in maternity hospitals at different times (Figure 1). *E. coli* isolates belong to various sequence types (STs), such as the globally spread ST1193, ST410, ST131, and ST617 or the occasionally isolated STs ST224, ST448, ST349, ST46, ST73, ST2491, and ST16617. Some of the STs, including ST410 (2), ST46 (2), and ST448 (2), were found in more than one hospital. ST1193 was isolated twice in HCY, in March and August, whereas some were isolated in only one hospital. The *Klebsiella* isolates were classified into two species and sequence types: two *K. pneumoniae* isolates belonging to ST1324 were isolated from two hospitals, whereas the two *K. quasipneumoniae* isolates belonging to ST489 were also isolated from two maternity units.

### Phylogenetic relationships of *E. coli* isolates

Core genome phylogenies were computed for the fifteen *E. coli* isolates, which were clustered according to sequence type (Figure 3).

**Figure 3.**
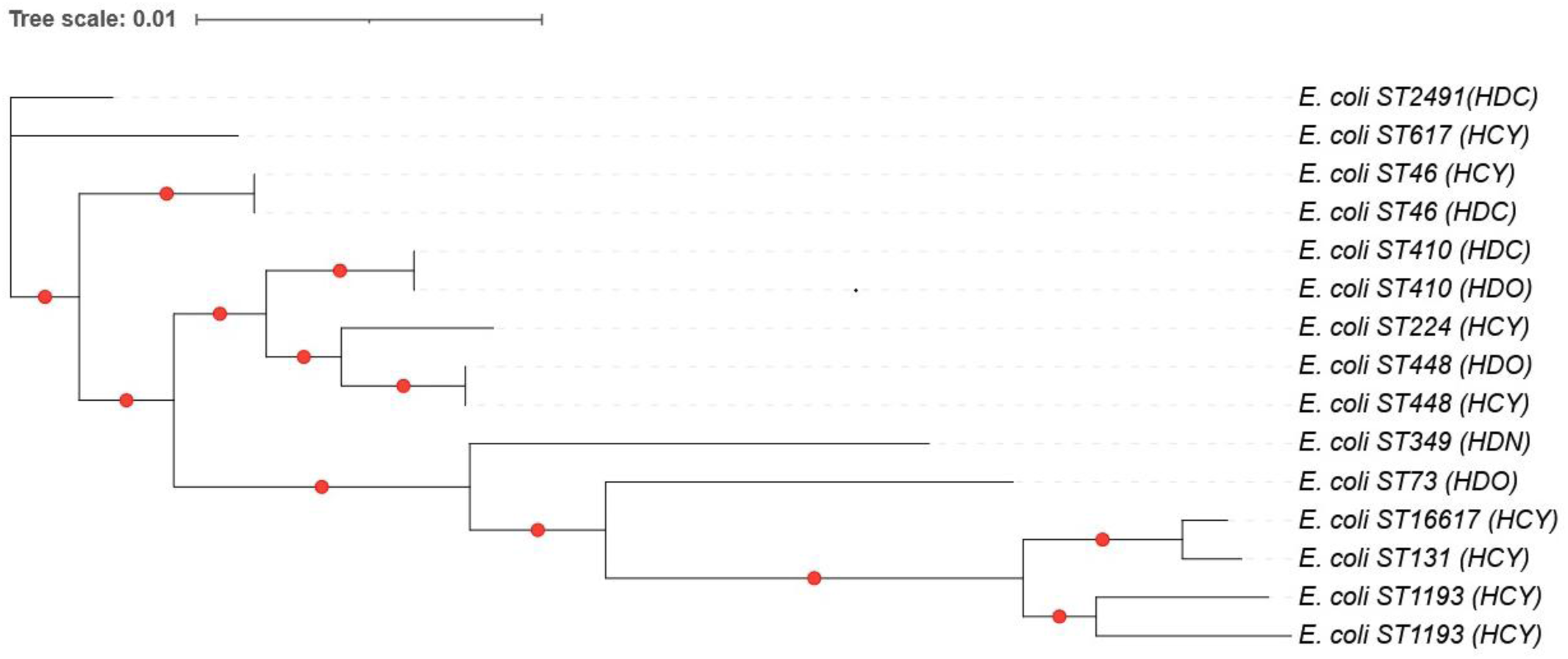
Core-genome maximum-likelihood phylogenetic tree of ESBL/carbapenemase producing *Escherichia coli* isolates recovered from in maternity ward environments in Yaoundé, Cameroon. The tree illustrates the genetic relatedness of environmental *E. coli* isolates and the distribution of high-risk lineages circulating across different maternity wards. Bootstrap values ≥ 70% are shown at key nodes.

The cluster of ST1193-related isolates was tight and contained two isolates from the same maternity ward (HCY). The ST73 isolate grouped closely with the ST1193 isolate, while the ST131 isolate formed a separate lineage but was still within the same major clade. A separate isolate of ST16617 fell in a separate lineage next to ST131. The ST410 isolates formed a large group, which came from two maternity wards.

The ST46 isolates came from two wards and formed a cluster. The two ST448 isolates from different wards were so closely related that they formed a subcluster. In contrast, isolates of the ST224, ST349, ST617 and ST2491 STs were located on distinct branches of the phylogenetic tree.

### Plasmid‒antibiotic resistance gene associations

WGS revealed that the ESBL-producing *E. coli* and *Klebsiella* isolates harboured highly diverse plasmid replicons and ARGs (Tables S2 and S3). Multiple replicons (plasmid incompatibility groups) associated with ARGs were identified. Replicons found in *Klebsiella* species included IncFII(K), IncFIB(K), IncL/M, and IncHI1B (pNDM-MAR), and each isolate had between two and three replicons each. These isolates were associated with various plasmid-associated ARGs, including ESBL genes (*bla*_CTX-M-15_, *bla*_SHV-11_, *bla*_SHV-18_, *bla*_OKP-B-6_), carbapenemase-associated genes (*bla*_OXA-2_), and additional resistance determinants targeting aminoglycosides, fluoroquinolones, sulfonamides, fosfomycin, chloramphenicol, and disinfectants (Figure 4).

**Figure 4.**
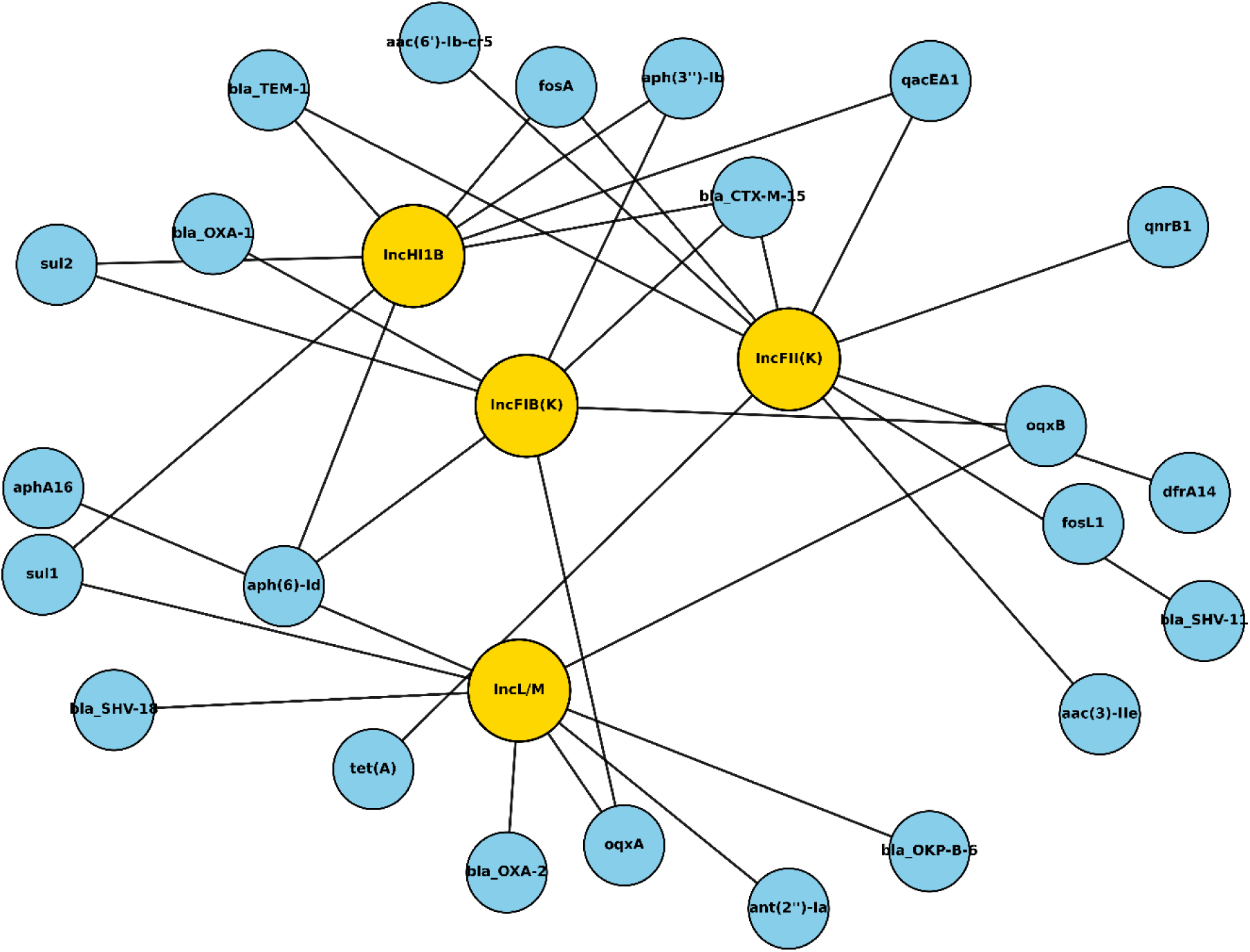
Plasmid-antibiotic resistance genes (ARGs) association network in ESBL/carbapenemase producing *Klebsiella* isolates recovered from maternity ward environmental surfaces in Yaoundé, Cameroon. Nodes represent plasmids or resistance genes, and edges indicate co-occurrence within the same isolate. The network highlights the central role of IncFII(K), IncFIB(K), IncL/M, and IncHI1B (pNDM-MAR) plasmids in carrying multiple β-lactam, aminoglycoside, fluoroquinolone, sulfonamide, fosfomycin, and disinfectant resistance determinants.

*E. coli* isolates carried a wide range of plasmid replicons, including IncFIA, IncFIB, IncFII, IncA/C2, IncL/M, IncX2, IncR, IncQ1, IncY, and several Col plasmids. Each isolate contained between one and 5 replicons. Epidemic plasmids such as IncFII (pAMA1167-NDM-5) and IncL/M (pOXA-48) were identified in various STs. Strong associations were found between IncF family plasmids (IncFIA, IncFIB, IncFII), IncA/C2 and IncL/M with ESBLs (*bla*_CTX-M-15_, *bla*_CTX-M-27_, *bla*_CTX-M-55_, *bla*_CTX-M-98_); AmpC β-lactamases (*bla*_CMY-2_, *bla*_CMY-6_); carbapenemases (*bla*_NDM-1_, *bla*_OXA-48_); aminoglycoside; macrolide, tetracycline, sulfonamide, trimethoprim, and chloramphenicol resistance genes; plasmid-mediated quinolone resistance determinants; and disinfectants, whereas IncX2, IncQ1, IncY, and Col plasmids presented more limited associations with specific subsets of resistance determinants (Figure 5).

**Figure 5.**
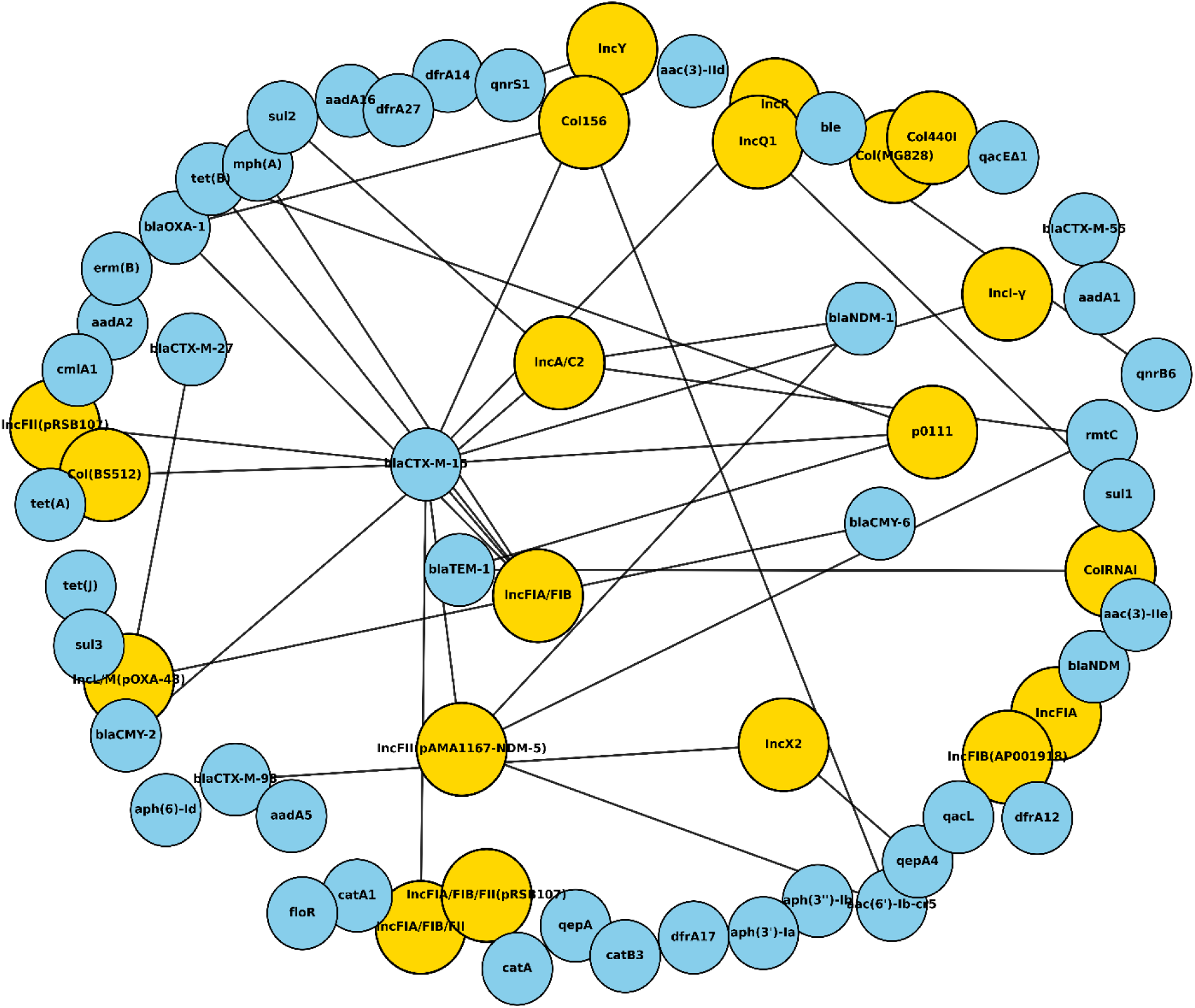
Plasmid-antibiotic resistance genes (ARGs) association network in ESBL/carbapenemase producing *Escherichia coli* isolates recovered from maternity ward environmental surfaces in Yaoundé, Cameroon. Plasmid replicon types are shown as yellow nodes and ARGs as blue nodes. Edges represent co-occurrence within the same isolate. The network highlights a dominant IncF plasmid backbone associated with extended-spectrum β-lactamase genes, alongside broad-host-range plasmids (IncA/C2 and IncL/M) carrying carbapenemase and multidrug resistance determinants.

### Virulence factor profiling

Diverse determinants of virulence were found in both isolates. The four ESBL *Klebsiella* isolates presented a highly conserved set of virulence genes (Table 1).

**Table 1.** Functional classification of virulence determinants identified in ESBL *Klebsiella* isolates recovered from maternity ward environmental surfaces in Yaoundé hospitals

| Functional category | Virulence determinants | *K. pneumoniae*<br>ST1324<br>(HDN) | *K. pneumoniae*<br>ST1324<br>(HCY) | *K. quasipneumoniae*<br>ST489 (HDO) | *K. quasipneumoniae*<br>ST489 (HCY) |
| --- | --- | --- | --- | --- | --- |
| Adhesins /<br>Biofilm<br>formation<br>(ECP operon) | ykgK/ecpR,<br>yagZ/ecpA,<br>yagY/ecpB,<br>yagX/ecpC,<br>yagW/ecpD,<br>yagV/ecpE | ✓ | ✓ | ✓ | ✓ |
| Iron<br>acquisition<br>(Enterobactin<br>system) | entA, entB,<br>fepC | ✓ | ✓ | ✓ | ✓ |
| Outer<br>membrane<br>proteins | ompA | ✓ | – | ✓ | ✓ |
| Motility /<br>flagellar<br>assembly | fliQ | – | ✓ | – | – |

Presence of virulence determinants was inferred from whole-genome sequencing data. ✓ indicates gene presence; – indicates absence.

The *ecp* operon genes, those involved in iron acquisition mediated by enterobactin (*entA*, *entB*, and *fepC*), adhesion or biofilm formation, were present in all the isolates. The outer membrane protein *OmpA* was present in three isolates; the flagella-related gene *fliQ* was detected in only one isolate (*K. pneumoniae* ST1324 isolated at HCY). The 15 isolates of *E. coli* presented a complex but organized virulence profile characterized by functions linked to persistence and host interaction (Table 2).

**Table 2.** Functional classification of virulence determinants identified in ESBL *Escherichia coli* isolates (n = 15) recovered from maternity ward environmental surfaces in Yaoundé hospitals

| Functional category | Representative virulence determinants | ST349 (HDN) | ST410 (HDO) | ST224 (HCY) | ST46 (HDC) | ST617 (HCY) | ST1661 7 (HCY) | ST131 (HCY) | ST448 (HDO) | ST119 3 (HCY-1) | ST119 3 (HCY-2) | ST410 (HDC) | ST249 1 (HDC) | ST46 (HCY) | ST73 (HDO) | ST448 (HCY) |
| --- | --- | --- | --- | --- | --- | --- | --- | --- | --- | --- | --- | --- | --- | --- | --- | --- |
| Adhesins / fimbriae / curli | fim, pap, sfa/foc, ecp, csg, upaH | ✓ | ✓ | ✓ | ✓ | ✓ | ✓ | ✓ | ✓ | ✓ | ✓ | ✓ | ✓ | ✓ | ✓ | ✓ |
| Iron acquisition / siderophores | ent, fep, chu, yersiniabactin, iuc/iut | ✓ | ✓ | ✓ | ✓ | ✓ | ✓ | ✓ | ✓ | ✓ | ✓ | ✓ | ✓ | ✓ | ✓ | ✓ |
| Capsule / serum resistance | kps, iss | ✓ | – | – | ✓ | – | – | ✓ | ✓ | ✓ | ✓ | – | – | – | ✓ | ✓ |
| Secretion systems | T2SS, T3SS, T6SS (esp, eiv, clpV) | ✓ | ✓ | ✓ | – | ✓ | ✓ | ✓ | ✓ | ✓ | ✓ | ✓ | ✓ | – | ✓ | ✓ |
| Toxins | hlyE, cnf1, astA, colibactin | ✓ | – | – | – | – | – | – | – | – | – | – | – | ✓ | ✓ | – |
| Motility / flagella | flagellar genes | ✓ | ✓ | ✓ | ✓ | – | ✓ | ✓ | ✓ | ✓ | ✓ | ✓ | ✓ | ✓ | ✓ | ✓ |
| Invasion / endothelial crossing | ibeB/C | ✓ | ✓ | ✓ | ✓ | ✓ | ✓ | ✓ | ✓ | ✓ | ✓ | ✓ | ✓ | ✓ | – | ✓ |
| Autotransporters / biofilm | sat, vat, biofilm-associated genes | – | ✓ | ✓ | – | ✓ | – | – | – | ✓ | ✓ | – | – | – | – | ✓ |

Presence of virulence determinants was inferred from whole-genome sequencing data. ✓ indicates presence of at least one gene within the functional category; – indicates absence.

Adhesiveness genes, biofilm secretion system-related genes, iron acquisition system genes, and motility genes represented the majority of the isolates. *The ebeB/C* genes involved in invasion and secretion systems were widely distributed. The high-virulence ExPEC clades associated with ST131, ST73, ST1193, and ST410 presented an organized virulence profile that included adhesiveness, iron uptake systems, and virulence genes linked to capsule/serum resistance. The genes linked to toxins and autotransporters were limited to a subset of isolates.

## Discussion

This study indicates that maternity ward environments in Yaounde, Cameroon, are colonized by a diverse collection of MDR Enterobacterales, including globally distributed high-risk lineages of *E. coli* and *Klebsiella* species.

All the isolates were recovered exclusively from environmental surfaces, with none being isolated from air samples, indicating surface-associated contamination. The major contamination was at the tertiary hospital (HCY), followed by lower recovery from the district hospitals (HDO, HDC, and HDN), suggesting a decrease in environmental contamination from tertiary to lower-level healthcare facilities.

This study combines phenotypic antimicrobial susceptibility testing with WGS to provide a detailed picture of ARGs, plasmid-mediated dissemination, virulence potential, and genetic relatedness of ESBL-producing *E. coli* and *Klebsiella* isolates circulating in these healthcare settings.

The isolation of these drug-resistant critical pathogens from high-contact maternity units, including intensive care, neonatology, delivery wards, hospitalization areas, and operating theatres, underscores the hospital environment as a significant reservoir for antibiotic resistance and the potential for infection in vulnerable patients. Previous studies reported that sinks, bed rails, work surfaces, and medical equipment can sustain long-term contamination with drug-resistant gram-negative bacteria, facilitating indirect transmission within healthcare facilities [13–15].

The recurrent detection of isolates over the sampling period and across multiple departments further supports persistent environmental contamination rather than periodic introduction.

Because neonates and postpartum mothers are very vulnerable and patient‒equipment interactions are frequent and intense, such contamination is especially concerning in maternity and neonatal units [16].

The detection of globally spread sequence types such as *K. pneumoniae* ST1324 and ST489 and *E. coli* ST131, ST1193, ST410, ST448, ST617, and ST73 points to multiple introductions and the ongoing circulation of high-risk clones rather than an isolated outbreak. These clones are known to have a powerful ability to acquire and disseminate ARGs through epidemic plasmids, and they are frequently reported in healthcare-associated infections [17,19].

As expected, the core-genome phylogenetic structure was influenced mainly by STs because STs designate stable evolutionary lineages and not transient epidemiological clusters [20]. As the wards’ metadata were not part of the tree-building process, the clusterings basically reflect genomic similarity rather than spatial proximity. In hospital genomic epidemiology, the fact that there is lineage-based clustering alone does not mean that transmission is ward restricted.

Relatedness must be carefully inferred. *E. coli* does not have a universally accepted SNP threshold for defining transmission; however, lower SNP distances are likely recent spreads, whereas high distances (e.g., around or even more than 100 core SNPs) are isolates that are incidentally and epidemiologically distinct [21,22]. Hence, genomic similarity must be considered together with temporal and epidemiological contexts. The fact that genetically similar strains occur in different wards and multiple departments may be due to the transmission of indirect pathogens through hospital staff, shared medical instruments, or patient transfer, which has been evidenced in hospital genomic investigations [21, 22]. On the other hand, wards may be different points of contact with large institutional or community reservoir-tracking samples [22]. Without longitudinal or patient flow information, it is impossible to determine the case from a cross-sectional study design [22].

These findings reinforce the need for a wide IPC intervention, including hand hygiene and environmental disinfection of highly touched surfaces [23,24]. Genomic surveillance with the use of next-generation sequencing technology can be integrated into existing AMR monitoring frameworks through the periodic sequencing of MDR isolates, which could complement phenotypic testing and facilitate the identification of high-risk clones at an early stage, in line with the recommendations of the WHO and Africa CDC [25,26]. A comparison of the phenotypic susceptibility results with the genomic resistance determinants revealed high overall concordance, with the concordance between genotype and phenotype exceeding 90% for β-lactams, fluoroquinolones, and aminoglycosides. No clear cases of phenotypic resistance lacking supporting genes were observed, and discrepancies were largely restricted to carbapenems. Resistance to penicillins, β-lactam/β-lactamase inhibitor combinations, and third-generation cephalosporins can be explained by the presence of ESBL genes, mainly *bla*_CTX-M-15_, which is still the most common gene globally and is strongly linked with successful ExPEC lineages [27, 28]. Resistance to cefoxitin in all the strains was associated with the presence of *bla*_CMY_ genes, which indicates plasmid-mediated AmpC production [29]. Fluoroquinolone resistance is completely accounted for by plasmid-mediated quinolone resistance genes and known lineage-related chromosomal mutations, particularly in strains ST131 and ST1193 [30, 31]. Aminoglycoside resistance can be explained by the presence of aminoglycoside-modifying enzyme-encoding genes.

Most of the mismatches were limited to carbapenems. In some cases, carbapenemase-positive isolates that were resistant to meropenem still presented variable susceptibility to imipenem. This finding reflects the differences in hydrolytic efficiency between enzyme classes and variants known from the literature, as well as being influenced by gene expression, plasmid copy number, and outer membrane permeability. The NDM enzymes belong to the class of metallo-β-lactamases, which are capable of hydrolysing both imipenem and meropenem very efficiently; nevertheless, phenotypic resistance levels may vary depending upon the bacterial background and regulatory factors [32, 33]. In contrast, OXA-48-like enzymes are typically weaker in carbapenem hydrolysis and may lead to borderline or heterogeneous resistance phenotypes, which in most cases result in increased resistance to meropenem compared with imipenem, especially when other permeability defects are absent [34, 35]. The phenotypic variability observed may be explained by the different enzyme classes and variants along with the different expression contexts rather than the assumption that a single carbapenemase gene behaves in the same way throughout.

The success of therapy depends to a very large extent on the phenotypic susceptibility pattern since the clinical efficacy is always determined by the measured MICs and not the simple presence of a resistance gene. On the other hand, detecting a gene remains essential for identifying carbapenemase producers that may exhibit borderline phenotypes and therefore be overlooked in routine diagnostics [36, 37]. This work endorses the use of both genomic and phenotypic methods as mutually supportive tools for identifying resistance mechanisms in multidrug-resistant Enterobacterales with the highest accuracy.

The accumulation of several aminoglycoside-modifying enzyme-encoding genes, such as *aac*, *aad*, and *aph* variants, which are frequently colocalized with β-lactam resistance genes on multidrug resistance plasmids, was also correlated with the high prevalence of aminoglycoside resistance. Colistin activity was maintained despite widespread resistance to several antibiotic classes, which is consistent with the lack of plasmid- mediated *mcr* genes and with the findings of several other studies [38]. However, reliance on colistin is still problematic in neonatal care because of its nephrotoxicity, neurotoxicity, and limited safety data.

However, in neonatal care, colistin is generally considered a last-resort option owing to its nephrotoxicity, neurotoxicity, and limited safety data, making reliance on it particularly concerning in maternity and neonatal settings.

Multiple diverse plasmid replicons have been identified, with multiple resistance determinants frequently cooccurring on mobile genetic elements. The plasmid families identified in this study, such as IncF, IncA/C2, IncL/M, and IncHI1B, are also frequently involved in the worldwide spread of ESBLs and carbapenemases [39–40].

Instead of being found only in clinical samples, their presence in hospital environment isolates emphasizes how the hospital setting can serve as a reservoir and an amplification niche for drug-resistant plasmids. The crucial role of particular plasmid backbones in connecting numerous resistance genes was further highlighted by plasmid-ARG association networks created for both *E. coli* and *Klebsiella* isolates. The discovery of efflux systems (such as *oqxAB*) and disinfectant resistance genes (such as *qacEΔ1*), which may decrease vulnerability to widely used biocides, was especially concerning. These characteristics may promote environmental persistence and compromise standard cleaning and disinfection procedures [41].

The isolates had a variety of virulence-associated factors in addition to antibiotic resistance genes. Toxins, adhesins, capsule-associated genes, invasion-associated genes (*ibeB/ibeC*) and iron acquisition systems are often found, especially in *E. coli* isolates from ExPEC lineages. Increased colonization, persistence and pathogenic potential, including neonatal infections, are linked to these characteristics [42–43]. Notably, given its documented role in host tissue damage and increased virulence, the discovery of a colibactin gene cluster in an ST73 isolate is especially concerning [36]. Concerns about the risks of maternal and newborn exposure in healthcare settings are heightened by the convergence of resistance, virulence, and environmental persistence traits, which implies that the environmental isolates from this study have the ability to colonize and possibly infect susceptible hosts.

## Conclusions

Our report shows that maternity ward environments in Yaounde are significant reservoirs of multidrug-resistant Enterobacterales, including high-risk, globally distributed lineages of *E. coli* and *Klebsiella* spp. The ability of the hospital setting to maintain and spread clinically significant resistance outside of patient-to-patient contact is highlighted by the discovery of epidemic ESBL- and carbapenemase-associated plasmid replicons across various STs. Because environmental contamination of frequently touched surfaces and water-associated sites can facilitate indirect transmission to highly vulnerable populations, these findings have direct implications for the health of mothers and newborns. Therefore, strengthening IPC measures, with a focus on targeted environmental decontamination and regular genomic surveillance, is crucial. Antimicrobial stewardship initiatives must be strengthened concurrently to lessen the selective pressure that promotes the upkeep and dissemination of these dangerous clones. This work emphasizes the importance of combining genomics and environmental sampling to guide context-specific interventions. These strategies are essential for protecting mothers and infants in healthcare settings with limited resources as well as for reducing the silent spread of MDR pathogens within and between maternity wards.

## Study limitations

The current study has certain limitations that need to be considered when the findings are interpreted. First, it was not possible to screen staff members in healthcare settings or include clinical samples from mothers or newborns; only environmental surfaces could be tested for the presence of antibiotic-resistant bacteria. As a result, establishing a direct link between an isolate’s environmental presence and any subsequent colonization or infection of patients or employees was impossible.

Second, the small number of isolates, particularly *Klebsiella* species, makes it difficult to assess the diversity of the isolates and differences between wards, as it is unlikely that all the ESBL-resistant isolates from the sampled wards were isolated in this study.

Third, a limited panel of antibiotics was used for antimicrobial susceptibility testing, and the minimum inhibitory concentration for each antibiotic was not assessed. This might have restricted the investigation of resistance levels, particularly with respect to colistin and carbapenems.

Fourth, long-read sequencing was not performed, which would have improved the assembly and examination of plasmid structures and links to resistance genes. Nevertheless, short-read sequencing has made it possible to thoroughly search for virulence factors, plasmids, and resistance genes.

Finally, systematic data on patient flow, cleaning procedures, antimicrobial use, and infection prevention strategies in maternity departments are lacking. This lack of context information made it more difficult to identify some of the contributing factors to environmental outbreaks.

Notwithstanding these drawbacks, the strength of combined phenotypic/genomic analysis is that it provides strong evidence of the diversity and presence of high-risk, multiresistant Enterobacterales in the maternity ward setting, prompting future investigations.

## Author contributions

GCB and GDA performed the data collection. GCB conducted the laboratory work. JUK and BPB conceptualized the study, acquired funding and managed the project. RN and FE participated in the conception of the study. BPB supervised all the students, performed the bioinformatic analysis and wrote the manuscript. NW advised the bioinformatic pipeline to analyse the data. JUK and MMCB contributed to the drafting and editing of the manuscript.

## Availability of data and materials

The assembled genomes of the 19 isolates have been deposited in the public database (NCBI GenBank database) under BioProject no. PRNJAXXXXX.

## Ethics approval

Ethical approval was obtained from the Centre Region Ethics Committee (00469/CRERSHC/2023). Additionally, administrative authorizations were also granted by the competent authorities of the investigated hospitals.

## Consent for publication

Not applicable.

## Data Availability

All data produced in the present study are available upon reasonable request to the authors

