## Supplementary Tables S1, S2, and S3 for "Environmental reservoirs of high-risk ESBL- and carbapenemase-producing *E. coli* and *Klebsiella* in maternity wards in Yaounde (Cameroon): Whole-genome sequencing and antimicrobial susceptibility studies"

**Table S1.** Antibiotic susceptibility profile of ESBL- *E. coli* and *Klebsiella* isolates from environmental surfaces of maternity wards in different hospitals in the city of Yaounde-Cameroon.

| | | **Isolates** | | | | | | | | | | | | | | | | | | |
|---|---|---|---|---|---|---|---|---|---|---|---|---|---|---|---|---|---|---|---|---|
| **Antibiotic family** | **Antibiotic** | *Escherichia coli ST1193 (HCY)* | *Escherichia coli ST410 (HDO)* | *Escherichia coli ST224 (HCY)* | *Escherichia coli ST448 (HCY)* | *Escherichia coli ST131 (HCY)* | *Escherichia coli ST1193 (HCY)* | *Escherichia coli ST617 (HCY)* | *Escherichia coli ST410 (HDC)* | *Escherichia coli ST448 (HDO)* | *Escherichia coli ST16617 (HCY)* | *Escherichia coli ST46 (HDC)* | *Escherichia coli ST349 (HDN)* | *Escherichia coli ST46 (HCY)* | *Escherichia coli ST2491 (HDC)* | *Escherichia coli ST73 (HDO)* | *Klebsiella quasipneumoniae ST489 (HCY)* | *Klebsiella pneumoniae ST1324 (HCY)* | *Klebsiella pneumoniae ST1324 (HDN)* | *Klebsiella quasipneumoniae ST489 (HDO)* |
| **β-lactams** | amoxicillin | R | R | R | R | R | R | R | R | R | R | R | R | R | R | R | R | R | R | R |
| | amoxicillin-clavulanic acid | R | R | R | R | R | R | R | R | R | R | R | R | R | R | R | R | R | R | R |
| | cefoxitin | R | R | R | R | R | R | R | R | R | R | R | R | R | R | R | R | R | R | R |
| | ceftriaxone | R | R | R | R | R | R | R | R | R | R | R | R | R | R | R | R | R | R | R |
| | ceftazidime | R | R | R | R | R | R | R | R | R | I | I | R | R | R | R | R | R | I | R |
| | cefotaxime | I | R | R | I | R | R | R | R | R | R | R | R | R | R | R | R | R | R | I |
| | aztreonam | S | R | R | R | R | R | R | R | R | R | S | R | R | R | R | R | S | R | R |
| | meropenem | R | R | R | R | R | R | R | R | R | R | R | R | R | R | R | R | R | R | R |
| | imipenem | S | S | S | S | S | S | S | S | S | S | S | S | S | S | S | S | S | S | S |
| **Aminosides** | gentamicin | R | I | R | R | R | R | R | R | I | S | S | R | R | S | S | S | R | R | S |

| | | | | | | | | | | | | | | | | | | | | |
|---|---|---|---|---|---|---|---|---|---|---|---|---|---|---|---|---|---|---|---|---|
| | tobramycin | R | R | R | I | R | R | R | R | I | R | R | R | I | R | R | R | I | R | R |
| **Fluoroquinolones** | ciprofloxacin | R | R | R | R | R | R | R | R | R | R | R | R | R | R | R | R | R | R | R |
| **Polymyxins** | colistin | S | S | S | S | S | S | S | S | S | S | S | S | S | S | S | S | S | S | S |

HCY: Yaoundé Central Hospital; HDCV: Cité Verte District Hospital; HDN: Nkolndongo District Hospital; HDO: Odza District Hospital.

R : Resistant, S : sensitive, I : intermediate

**Table S2.** Summary of plasmids and antibiotic resistance genes identified in four *Klebsiella* isolates recovered from maternity ward environmental surfaces in Yaoundé hospitals

| Species / ST | Hospital | Plasmids detected | Antibiotic resistance genes |
|---|---|---|---|
| *K. pneumoniae* ST1324 | HDN | IncFII(K); IncFIB(K) (Kpn3-type) | $^{bla}$CTX-M-15, $^{bla}$TEM-1, $^{bla}$SHV-11, $^{bla}$OXA-1, *fosL1, dfrA14, qnrB1, tet(A), aph(6)-Id, aph(3'')-Ib, sul2, qacEdelta1, fosA, aac(6')-Ib-cr5, aac(3)-IIe, oqxA, oqxB19, fosA.* |
| *K. quasipneumoniae* ST489 | HDO | IncL/M (pMU407); IncFII(K); IncFIB(K) | $^{bla}$OXA-2, $^{bla}$SHV-18, $^{bla}$OKP-B-6, *aphA16, ant(2'')-Ia, sul1, qacEdelta1, oqxA, oqxB, fosA.* |
| *K. pneumoniae* ST1324 | HCY | IncHI1B (pNDM-MAR); IncFIB(K) (Kpn3-type); IncFII(K) | $^{bla}$CTX-M-15, $^{bla}$TEM-1, $^{bla}$SHV-11, *aph(6)-Id, aph(3'')-Ib, sul2, tet(A), qnrB1, dfrA14, aac(3)-IIe, aac(6')-Ib-cr5, blaOXA-1, fosA, qacEdelta1, sul1, oqxA, fosA.* |
| *K. quasipneumoniae* ST489 | HCY | IncL/M (pMU407); IncFII(K); IncFIB(K) (Kpn3-type) | $^{bla}$SHV-18, $^{bla}$OKP-B-6, $^{bla}$OXA-2, *ant(2'')-Ia, aphA16, qacEdelta1, sul1, oqxB, oqxA, fosA.* |

*HCY*: Yaoundé Central Hospital; *HCV*: Cité verte district hospital; *HDN*: Nkolndongo district hospital; *HDO*: Odza district hospital

**Table S3.** Summary of plasmids and antibiotic resistance genes (ARGs) identified in fifteen ESBL *E. coli* sequence types (ST) recovered from maternity ward environmental surfaces in Yaoundé hospitals

| ST | Hospital | Plasmids detected | Key Antibiotic Resistance Genes (ARGs) |
|---|---|---|---|
| ST349 | HDN | p0111 | $^{bla}$CTX-M-15, $^{bla}$OXA-1, $^{bla}$TEM-1, *mph(A), sul1, qacEdelta1, aadA2, dfrA12, qepA4, aadA1, tet(B).* |
| ST410 | HDO | IncFII(pAMA1167-NDM-5), IncFIA/FIB, ColRNAI, Col(BS512), Col(MG828) | $^{bla}$CMY-2, $^{bla}$CTX-M-15, $^{bla}$OXA-1, $^{bla}$TEM-1, *mph(A), dfrA12, aadA2, qacEdelta1, sul1, tet(B), aac(6')-Ib-cr5.* |
| ST224 | HCY | IncR, Col440I | $^{bla}$TEM-1, $^{bla}$CTX-M-15, $^{bla}$SCO-1, *aac(3)-IIe, dfrA12, aadA2, cmlA1, aadA1, qacL, sul3, aac(6')-Ib-cr5, arr-3, dfrA27, aadA16, qacEdelta1, sul1, qnrB6, tet(A)* |
| ST46 | HDC | IncX2, IncFIA/FIB, IncQ1, Col156 | $^{bla}$TEM-1, $^{bla}$CTX-M-98, $^{bla}$OXA-1, *aadA1, aph(3')-Ia, sul2, aph(3'')-Ib, aph(6)-Id, tet(B), qepA4.* |
| ST617 | HCY | IncFIA/FIB, IncFII(pAMA1167-NDM-5) | $^{bla}$CTX-M-15, $^{bla}$TEM-1, *aac(3)-IId, catA1, qepA, dfrA12, aadA2, qacEdelta1, sul1, tet(B), mph(A).* |
| ST16617 | HCY | IncFIB(AP001918), IncFIA, IncI-γ, IncFII(pRSB107) | $^{bla}$CTX-M-15, $^{bla}$OXA-1, *aac(6')-Ib-cr5, sul2, aph(3'')-Ib, aph(6)-Id, tet(A), mph(A), sul1, catB3, qacEdelta1, aadA5, dfrA17, aac(3)-IIe* |
| ST131 | HCY | IncL/M(pOXA-48), IncA/C2, IncFIA/FIB/FII, Col156 | $^{bla}$CMY-6, $^{bla}$CTX-M-27, *rmtC, tet(J), catA, mph(A), dfrA17, aadA5, qacEdelta1, sul1, tet(A), aph(6)-Id, aph(3'')-Ib, sul2, erm(B).* |
| ST448 | HDO | IncFII(pAMA1167-NDM-5), IncA/C2, IncFIA/FIB, p0111, IncQ1 | $^{bla}$OXA-1, $^{bla}$CMY-2, $^{bla}$TEM-1, $^{bla}$NDM-1, $^{bla}$CTX-M-15, *tet(J), catA, dfrA17, aadA5, qacEdelta1, sul1, rmtC, ble, aac(3)-IId, sul2, aph(3'')-Ib, aph(6)-Id, mph(A), tet(B), aac(6')-Ib-cr5.* |
| ST1193 | HCY | IncFIA, Col(BS512) | $^{bla}$CTX-M-15, $^{bla}$OXA-1, *aac(6')-Ib-cr5, aac(3)-IIe.* |
| ST1193 | HCY | IncA/C2, IncFIA/FIB, IncQ1, Col(BS512), Col156 | $^{bla}$OXA-1, $^{bla}$NDM-1, $^{bla}$CTX-M-15, *tet(J), aac(6')-Ib-cr5, aac(3)-IIe, catA, ble, rmtC, tet(B), sul2, aph(3'')-Ib, aph(6)-Id, dfrA17, sul1, qacEdelta1.* |
| ST410 | HDC | IncFII(pAMA1167-NDM-5), IncFIA/FIB, IncQ1, Col plasmids | $^{bla}$CMY-2, $^{bla}$CTX-M-55, $^{bla}$NDM, $^{bla}$OXA-1, $^{bla}$TEM-1, *sul2, aph(3'')-Ib, aph(6)-Id, tet(A), floR, qacEdelta1, sul1, ble, aac(3)-IId, tet(B), dfrA17, aac(6')-Ib-cr5.* |
| ST2491 | HDC | IncY | $^{bla}$CTX-M-15, $^{bla}$TEM-1, *qnrS1, aph(6)-Id, aph(3'')-Ib, sul2, tet(A), dfrA14* |
| ST46 | HCY | IncX2, IncFIA/FIB, Col156 | $^{bla}$TEM, $^{bla}$CTX-M-98, $^{bla}$OXA-1, *aadA1, aph(3')-Ia, tet(B), qepA4, aph(3'')-Ib, aph(6)-Id.* |
| ST73 | HDO | IncFIA/FIB/FII(pRSB107), Col156 | $^{bla}$OXA-1, $^{bla}$CTX-M-15, *sul2, aph(3'')-Ib, aph(6)-Id, tet(A), sul1, qacEdelta, aadA5, dfrA17, aac(6')-Ib-cr5.* |
| ST448 | HCY | p0111, IncA/C2, IncFIA/FIB/FII(pAMA1167-NDM-5), IncQ1 | $^{bla}$CMY-2, $^{bla}$TEM-1, $^{bla}$CTX-M-15, $^{bla}$OXA-1, *tet(B), mph(A), aac(3)-IId, sul2, aph(3'')-Ib, aph(6)-Id, dfrA17, aadA5, qacEdelta1, sul1, aac(6')-Ib-cr5.* |

*HCY*: Yaoundé Central Hospital; *HCV*: Cité verte district hospital; *HDN*: Nkolndongo district hospital; *HDO*: Odza district hospital
